# Fear of COVID-19 after vaccination dissemination and its relationship with multidimensional health literacy among patients on maintenance haemodialysis

**DOI:** 10.1101/2024.10.19.24315821

**Authors:** Atsuro Kawaji, Ryohei Inanaga, Mamiko Ukai, Tetsuro Aita, Yusuke Kanakubo, Takumi Toishi, Masatoshi Matsunami, Tatsunori Toida, Yu Munakata, Tadao Okada, Tomo Suzuki, Noriaki Kurita

## Abstract

**Background and hypothesis.:** The increased anxiety owing to the COVID-19 pandemic has been suggested to contribute to unhealthy lifestyles and depression in patients undergoing haemodialysis (HD). Therefore, this study aimed to evaluate the degree of fear of COVID-19 after vaccination dissemination and the independent impact of high-order health literacy (HL) on fear, which have not been adequately investigated.

**Methods.:** This multicentre cross-sectional study, conducted in 2022, after the widespread availability of the COVID-19 vaccination in Japan, included adults undergoing in-centre HD. Multidimensional HL was measured using the 14-item Functional, Communicative, and Critical Health Literacy Scale. Fear of COVID-19 was measured using the 7-item Japanese version of the Fear of COVID-19 Scale. COVID-19 fear scores in patients with HD were compared with scores of adults in April 2020 (the beginning of the pandemic) using an unpaired t-test. The association between multidimensional HL and COVID-19 fear scores was estimated using a multivariable-adjusted general linear model.

**Results.:** A total of 446 patients were analysed, of whom 431 (97%) and nine (2%) received three and two doses of vaccination, respectively. Their COVID-19 fear scores were significantly lower than those of the general population at the beginning of the pandemic (p < 0.001; mean difference -4.4 [95% confidence interval (CI): -5.1 – -3.7]; standardised effect size [ES] 0.77). Higher functional HL was associated with less fear (per 1-pt higher: -2.8 [95% CI: -1.7 – -0.3]; standardised ES -0.51), whereas higher critical HL was associated with greater fear (per 1-pt higher: 3.2 [95% CI: 0.7 – 3.0]; standardised ES 0.80). Communicative HL was not associated with fear.

**Conclusion.:** Patients’ fear of low-functional HL, despite widespread vaccination, can be reduced by providing health information in an easy-to-understand manner. Thus, the fear of sceptics owing to excessive critical HL and honest explanations by healthcare providers may be important.

**Key learning points What was known:** Heightened anxiety stemming from the COVID-19 pandemic exacerbates unhealthy lifestyles and depression, particularly in patients undergoing haemodialysis.

Health literacy plays a crucial role in individuals with kidney disease and may mitigate anxiety.

Comprehensive data on whether multidimensional health literacy (functional, critical, and communicative) independently correlates with fear of COVID-19, particularly in the context of patients undergoing haemodialysis are lacking.

**This study adds:** COVID-19 fear scores were notably lower in patients undergoing haemodialysis who received the COVID-19 vaccination than in the general population at the onset of the pandemic.

Elevated functional health literacy correlated with reduced fear, whereas higher critical health literacy was linked to increased fear.

**Potential impact:** Fear among individuals with low functional health literacy can be alleviated by delivering health information in a clear and accessible manner, whereas transparent and honest communication from healthcare providers is crucial for patients with heightened fear owing to critical health literacy, who may be sceptical of accurate information.

## Introduction

Fear of the COVID-19 pandemic causes considerable anxiety disorders, especially among patients with chronic kidney disease (CKD) who are at high risk of severe disease. In particular, patients undergoing haemodialysis who are required to share a specific space and spent time in a dialysis unit would have felt strong fear during epidemic periods.

Although strong fear and anxiety may have increased adherence to vaccination and wearing of masks in public places, they have also been suggested to have contributed to unhealthy lifestyle habits, weight gain, isolation, and depression among patients with CKD (1,2).

Considering the potentially increased risk of all-cause mortality and 1-year hospitalisation rates associated with anxiety symptoms (3), the identification of related factors and implementation of measures to mitigate fear is clinically important. Health literacy (HL), or the ability to understand, discern, or use appropriate information, may help mitigate anxiety because patients with CKD are likely to be confused by inconsistent information about COVID-19 (2). Consequently, HL was viewed as a social vaccine that empowered people to protect their own and high-risk populations’ health during the COVID-19 pandemic (4).

Given that many patients with CKD have low HL (5), examining the impact of HL on fear of COVID-19 is important; however, it remains poorly studied.

Although one study showed that low HL was associated with high anxiety among patients with preserved CKD (1), only “conventional” HL was assessed. Recently, however, the importance of assessing multidimensional HL has increased (6,7). That is, in addition to the most basic functional HL, such as reading and writing, communicative HL (i.e., the ability to extract information, infer meaning, and apply information to changing situations) and critical HL (i.e., the ability to critically analyse information and control current situations) are also gaining attention (6). One small study showed that higher levels of each of these multidimensional HL dimensions correlated with lower levels of COVID-19 anxiety among patients undergoing haemodialysis. However, the study failed to examine how individual dimensions independently affected anxiety about COVID-19, even though each of the HL dimensions was correlate with the others (8). Therefore, identifying the dimensions of HL that independently influence the fear of COVID-19 in patients undergoing haemodialysis would be useful in developing support tailored to their individual health literacy.

Furthermore, the potential to identify and address patients with excessive fear persists despite the spread of effective measures against COVID-19, such as vaccines.

Therefore, we aimed to quantify the degree of fear of COVID-19 among patients undergoing maintenance haemodialysis after vaccine dissemination and evaluate whether multidimensional HL (functional, critical, and communicative) was independently associated with fear of COVID-19.

## Materials and Methods Study Design and Participants

This multicentre cross-sectional study was conducted at six medical institutions providing outpatient haemodialysis: Kameda Medical Center (Chiba), Kameda Family Clinic Tateyama (Chiba), Awa Regional Medical Center (Chiba), Munakata Clinic (Chiba), Chikuseikai Munakata Clinic (Tokyo), and Shinyurigaoka General Hospital (Kanagawa). The participants were adult patients with End-stage kidney disease (ESKD) who regularly visited the participating facilities and received either haemodialysis or a hybrid treatment of haemodialysis and peritoneal dialysis. Patients deemed by medical staff as unable to respond to the study questionnaire by themselves were excluded from the study. A paper- based questionnaire was distributed at each facility, and the participants were asked to complete the questionnaire at home and return it to the medical staff without identifying the content. The respondents were provided with a 500-yen gift card as an incentive. In addition, the attending physician extracted dialysis-related patient data from medical records. Data were collected between June 2022 and February 2023, after the spread of the COVID-19 vaccine in Japan. This study was conducted in accordance with the Declaration of Helsinki and approved by the Institutional Review Board of Fukushima Medical University (approval number: ippan2021-292). All participants were provided with information about the study in advance, and written informed consent was obtained from all participants.

## Measures

### Outcome: Fear of COVID-19

The main outcome, fear of COVID-19, was measured using the Japanese version of the COVID-19 Scale (9,10). This scale consists of seven items and responses are scored on a 5-point Likert scale ranging from 1 (strongly disagree) to 5 (strongly agree). The total score was calculated by adding the scores for each item (7–35 points), with a higher score indicating a stronger fear of COVID-19. The Japanese version of the scale has been validated in the general population and has been confirmed to have internal consistency reliability (α = 0.87) and validity (9).

### Exposure: multidimensional health literacy

The main exposure, multidimensional HL, was measured using the Functional, Communicative, and Critical Health Literacy (FCCHL) scale (11). This self-report scale consists of 14 items and can be scored on three subscales: Functional HL (five items), Communicative HL (five items), and Critical HL (four items). Responses to each item were scored on a four- point Likert scale ranging from 1 (never) to 4 (often). The scores were reversed when functional HL was measured. The mean score of the corresponding items was used as the scale score for each of the three subscales, with higher scores indicating higher HL levels of health literacy. This scale has previously shown high internal consistency for functional, communicative, and critical HL (alpha coefficients of 0.84, 0.77, and 0.65, respectively) in Japanese patients with type 2 diabetes, demonstrating its construct validity and reliability (11).

### Measurement of Covariates

Based on the literature and expert knowledge, potential confounding factors that could affect health literacy and fear of COVID-19 were identified. The covariates included age, sex, duration of dialysis, smoking history, educational level, household income, and comorbidities. Comorbidities were scored based on the modified Charlson Comorbidity Index (CCI) for patients undergoing dialysis (12,13).

### Statistical Analyses

Statistical analyses were performed using Stata/SE, version 17 (StataCorp, College Station, TX, USA). Patient characteristics are presented as median and interquartile range (IQR) for continuous variables and as frequencies and percentages for categorical variables. Differences between the fear of COVID-19 scores in the present study and those of 450 Japanese adults in April 2020 (i.e. the beginning of the COVID-19 pandemic) reported in the development paper were compared using an unpaired t-test (9). The association between the aforementioned covariates and each domain of multidimensional HL was estimated using general linear models. Subsequently, the association between multidimensional HL and fear of COVID-19 was estimated using a general linear model. The above-mentioned covariates were adjusted. All models were estimated using robust standard errors to address heteroscedasticity. We assumed that the data analysed were missing at random, and imputed the missing data using multiple imputations with chained equations to perform 20 imputations (14). A two-sided significance level of p < 0.05 was used for each analysis.

## Results

### Study flow and patient characteristics

Among the 651 patients undergoing haemodialysis at six facilities, 484 (74.3%) participated in the study. After excluding 38 cases with missing data on the fear of COVID- 19, 446 cases were analysed. Patient characteristics are shown in Table 1. The median age of the patients was 69 years (IQR 63–78), and 157 (35.2%) patients were women. A total of 61.6% patients had graduated from high school and 79.8% patients had a household income of < 5 million yen. A total of 431 (96%) and nine (2%) patients received three and two doses of the COVID-19 vaccine, respectively. The median scores for the functional, communicative, and critical HL subdomains were 3.2 (IQR 2.6–3.8), 2.8 (IQR 2.2–3.2), and 2.5 (IQR 2.0–3.0), respectively.

**Table 1.** Patient characteristics† (N = 446)

|  |  | Missing,<br>n |
| --- | --- | --- |
| Age, years | 69.4 (63.0–78.0) | 0 |
| Women, n (%) | 157 (35.2) | 0 |
| Kidney disease, n (%) |  | 0 |
| Diabetic nephropathy | 166 (37.2) |  |
| Chronic glomerulonephritis | 102 (22.9) |  |
| Nephrosclerosis | 61 (13.7) |  |
| Polycystic kidney disease | 14 (3.1) |  |
| Others | 68 (15.3) |  |
| Unknown | 35 (7.9) |  |
| Dialysis duration, year | 5.8 (2.9–10.6) | 0 |
| Comorbidities, n (%) |  |  |
| Cardiovascular disease | 187 (42.3) | 4 |
| Depression | 10 (2.2) | 1 |
| Diabetes | 192 (43.3) | 3 |
| Malignancy | 47 (10.6) | 1 |
| Leukaemia | 2 (0.5) | 1 |
| Malignant lymphoma | 5 (1.1) | 1 |
| Dementia | 19 (4.3) | 2 |
| Chronic liver disease | 25 (5.6) | 1 |
| Peptic ulcer | 33 (7.4) | 0 |
| Chronic lung disease | 11 (2.5) | 3 |
| Collagen disease | 11 (2.5) | 2 |
| modified Charlson score, points | 1.0 (0–2) | 0 |
| Smoking, n (%) |  | 21 |
| Never | 232 (54.6) |  |
| Ever | 145 (34.1) |  |
| Current | 48 (11.3) |  |
| Household income, n (%) |  | 35 |
| < 1,000,000 yen | 37 (9.0) |  |
| 1,000,000– < 5,000,000 yen | 291 (70.8) |  |
| 5,000,000– < 10,000,000 yen | 69 (16.8) |  |
| > 10,000,000 yen | 14 (3.4) |  |
| Education level, n (%) |  | 19 |
| Junior high school graduate or less | 102 (23.9) |  |
| High school graduate | 263 (61.6) |  |
| University/Graduate school graduate | 57 (13.3) |  |
| Others | 5 (1.2) |  |
| COVID-19 vaccination, n (%) |  | 0 |
| Unvaccinated | 1 (0.2) |  |
| Once | 5 (1.1) |  |
| Twice | 9 (2.0) |  |
| Thrice | 431 (96.6) |  |
| Functional HL, per 1-pt higher | 3.2 (2.6–3.8) | 14 |
| Communicative HL, per 1-pt higher | 2.8 (2.2–3.2) | 24 |
| Critical HL, per 1-pt higher | 2.5 (2.0–3.0) | 13 |

<sup>†</sup>Continuous variables are summarised as median (interquartile range). Categorical variables are presented as frequencies and proportions (parentheses).
3 According to the "Comprehensive Survey of Living Conditions" conducted by the Ministry of 4 Health, Labour and Welfare in 2022, the percentages of household income < 1 million yen, 5 1–3 million yen, 3–5 million yen, 5–10 million yen, and >10 million yen were 7%, 28%, 23%, 6 30%, and 13%, respectively.

### Comparison of the fear of COVID-19 between study participants and the general population in the early phase of the pandemic

The mean and standard deviation of the fear of COVID-19 scores among the participants was 16.9 and 5.7 points, which was significantly lower than the score of the general population in the early phase of the pandemic (p < 0.001; mean difference -4.4 [95% confidence interval (CI): -5.1 to -3.7]; standardised effect size (ES) 0.77). (9) (Figure 1):

**Figure 1.**
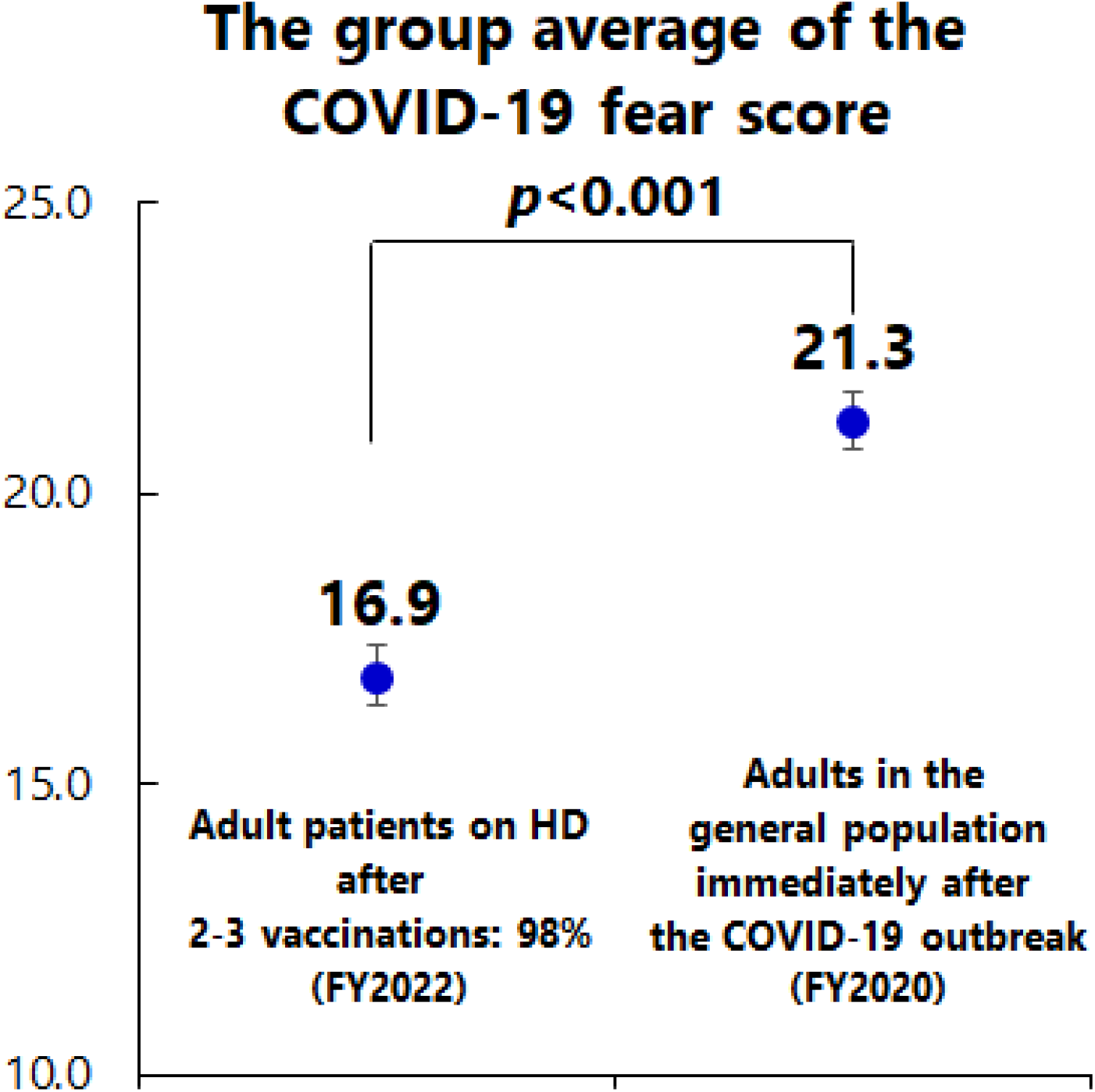
Comparison of group mean of COVID-19 fear scores Using the mean and standard deviation of fear scores for COVID-19 in the present study and the corresponding values of the scores from 450 Japanese adults in April 2020 (i.e. at the beginning of the COVID-19 outbreak), as reported in a development paper, these groups were compared using an unpaired t-test. Error bars indicate 95% confidence intervals. The mean difference for this population relative to the Japanese adults was -4.38 points (95% confidence interval -5.11 – -3.65).

### Association between HL subdomains and patient characteristics

Table 2 shows the association between the HL subdomains and covariates. Functional HL was positively associated with dialysis duration (per 1 year higher: 0.01 [95% CI: 0.002 – 0.02]) and education (especially high school, university, and other) (vs. junior high school or lower: 0.37 [95% CI: 0.16 – 0.58], 0.64 [95% CI: 0.39 – 0.89], 0.87 [95% CI: 0.27 – 1.46]) but negatively associated with comorbidity score (per 1 point higher: -0.66 [95% CI: -0.11 – -0.02], respectively). Communicative HL was positively associated with educational level (especially university graduate, high school, other) (vs. junior high school or lower: 0.50 [95% CI: 0.27 – 0.74], 0.17 [95% CI: 0.0004 – 0.34], 0.63 [95% CI: 0.07 – 1.12], respectively) and was negatively associated with age (per 1 year higher: -0.07 [95% CI: -0.12 – -0.002]) and comorbidity score (per 1 point higher: -0.05 [95% CI: -0.08 – -0.01]). Critical HL was positively associated with educational level (especially university graduate, other) (vs. junior high school or lower: 0.46 [95% CI: 0.21 – 0.71], 0.58 [95% CI: 0.07 – 2.20]) but negatively associated with comorbidity score (per 1-point higher: -0.04 [95% CI: -0.07 – - 0.006]).

**Table 2.**
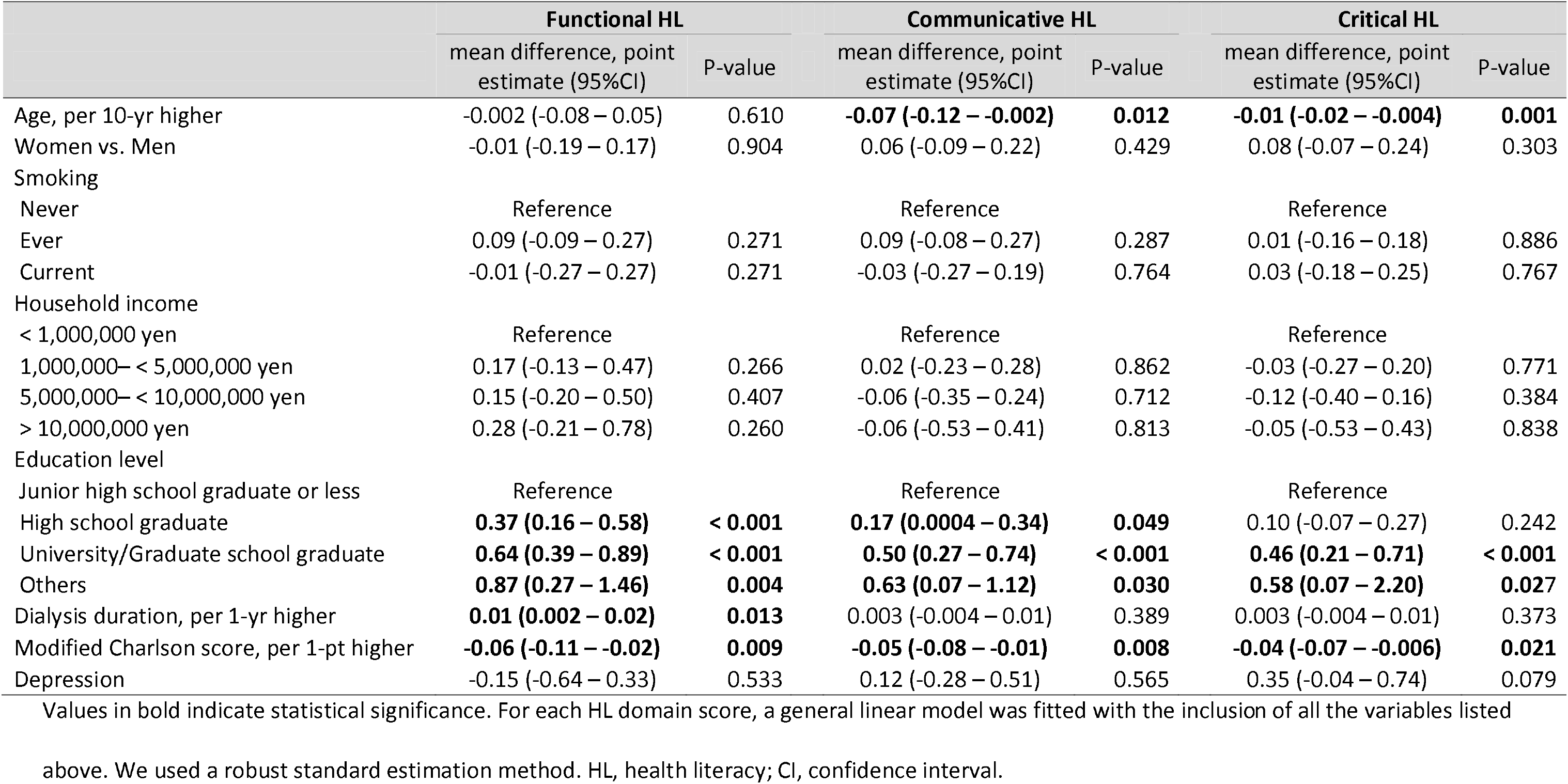
Associations of health literacy domains with covariates† (N = 446)

### Association between fear of COVID-19 and multidimensional HL

Table 3 shows the associations among fear of COVID-19 and HL subdomains, and covariates. Fear of COVID-19 was negatively associated with functional HL (per 1-point higher: -0.99 [95% CI: -1.72 – -0.26]) and household income (especially > 5 million yen – < 10 million yen) (reference: < 1 million yen: -3.01 [95% CI: -5.41 – -0.61]). In contrast, higher critical HL (per 1-point higher: 1.85 [95% CI: 0.74 – 2.97]) and women (vs. men; 1.98 [95% CI: 0.73 – 3.23]) were positively associated with fear of COVID-19. Communicative HL was not associated with fear.

**Table 3.** Associations of COVID-19 fear score with health literacy and covariates† (N = 446)

|  | Corresponding<br>standardised<br>ES | mean difference,<br>point estimate<br>(95%CI) | P-value |
| --- | --- | --- | --- |
| **Functional HL, per 1-pt higher** | **-0.18** | **-0.99 (-1.72 – -0.26)** | **0.008** |
| Communicative HL, per 1-pt higher | -0.10 | -0.59 (-1.69 – 0.51) | 0.291 |
| **Critical HL, per 1-pt higher** | **0.34** | **1.85 (0.74 – 2.97)** | **0.001** |
| Age, per 10-yr higher | 0.006 | 0.03 (-0.01 – 0.08) | 0.160 |
| **Women vs. Men** | **0.37** | **1.98 (0.73 – 3.23)** | **0.002** |
| Smoking |  |  |  |
| Never |  | Reference |  |
| Ever | -0.15 | -0.79 (-2.06 – 0.47) | 0.238 |
| Current | -0.08 | -0.42 (-2.22 – 1.39) | 0.649 |
| Household income |  |  |  |
| < 1,000,000 yen |  | Reference |  |
| 1,000,000– < 5,000,000 yen | -0.25 | -1.37 (-3.65 – 0.93) | 0.233 |
| **5,000,000– < 10,000,000 yen** | **-0.56** | **-3.01 (-5.41 – -0.61)** | **0.014** |
| > 10,000,000 yen | **-0.61** | **-3.29 (-6.49 – 0.10)** | **0.043** |
| Education level |  |  |  |
| Junior high school graduate or less |  | Reference |  |
| High school graduate | -0.11 | -0.57 (-1.97 – 0.85) | 0.420 |
| University/Graduate school<br>graduate | -0.30 | -1.60 (-3.42 – 0.22) | 0.084 |
| Others | 0.62 | 3.36 (-1.55 – 8.28) | 0.179 |
| Dialysis duration, per 1-yr higher | 0.006 | -0.03 (-0.04 – 0.10) | 0.367 |
| Modified Charlson score, per 1-pt<br>higher | -0.02 | -0.11 (-0.40 – 0.18) | 0.450 |
| Depression | 0.07 | 0.39 (-3.03 – 3.81) | 0.825 |

Values in bold indicate statistical significance. The general linear model was fitted with the
inclusion of all the variables listed above. We used a robust standard estimation method. To
calculate the corresponding standardised effect size (Cohen *d*), the point estimate was
divided by the SD of the COVID-19 fear scores to calculate the corresponding standardised
effect size (Cohen's *d*). CI, confidence interval; ES, effect size; HL, health literacy.

## Discussion

Our results indicated that fear of COVID-19 among adult Japanese patients on haemodialysis was significantly lower than that of the general population in the early phase of the pandemic after the widespread distribution of the COVID-19 vaccine. However, the findings that low functional HL and high critical HL were associated with increased fear of COVID-19 highlight the need for measures tailored to individual HL even after the dissemination of the vaccine.

Although previous research on the level of anxiety after the widespread use of COVID-19 vaccines among patients on dialysis is scarce, findings on the relationship between multidimensional HL and COVID-19-related anxiety are similar to those found in the non-dialysis population. Older age and a history of diabetes, heart disease, or lung disease were associated with an increased likelihood of having anxiety related to COVID-19 in a study conducted on the general population in the UK during the early period of the COVID-19 pandemic (March 2020) (15). There are a few possible reasons why our patients with the same serious illness as those mentioned above were less anxious about COVID-19 than the general population before the pandemic. First, the high vaccination rate (98%) and alleviation of uncertainty regarding the pathogenesis and prevention of COVID-19 in the early phase of the pandemic may have reduced anxiety in our study participants. Similarly, although patients on haemodialysis were more anxious than those on peritoneal dialysis before the vaccination programme in a study comparing patients on haemodialysis and peritoneal dialysis in the Netherlands, no such difference was observed after the programme (16). However, as noted in a previous synthesised qualitative study, the uncertainty about healthcare experienced during the pandemic, such as the disruption of care, confusion owing to a lack of reliable information, and scepticism about vaccines, undoubtedly contributed to anxiety among patients on dialysis (2).

Another explanation is that the mental health of patients on dialysis was resilient to or unaffected by the COVID-19 pandemic. This is consistent with a previous study wherein mental health was not affected during the pandemic in Dutch patients with CKD or in those on dialysis who did not develop COVID-19 (17). Additionally, the limited impact of social distancing measures compared to that in the general population (18), the strengthening of family ties, and the sense of safety towards healthcare institutions owing to strict preventative measures (2,18) may have helped alleviate anxiety.

A U.S. study on fear of COVID-19 linked to low health literacy among patients with non-dialysis-dependent CKD only examined functional HL (1). Whereas, all three types of HLs (functional, communicative, and critical) showed a negative correlation (i.e. low health literacy was associated with a high fear of COVID-19) with fear of COVID-19 among patients on haemodialysis (8); however, this study did not adjust for confounding factors. In contrast, as shown in the current study, differences owing to confounding factors such as age, sex, education history, and comorbidities were not addressed. Additionally, although our finding of higher critical HL along with a greater fear of COVID-19 contradicts the Korean study; it is consistent with a survey of the Japanese general population (19). The fear of COVID-19 among groups with high HL in that study was also directed toward government and social policies. (19) Moreover, higher critical HL has been suggested to be associated with worse medication adherence (particularly in terms of behavioural aspects) among patients on haemodialysis, mainly owing to a loss of trust in physicians (7). Additionally, patients with high critical HL may have gathered too much information about the pandemic resulting in unnecessary scepticism about the gathered information, including the best information provided by the government and physicians at the time.

This study has several clinical implications for dialysis providers. First, communicating the correct medical information about COVID-19 using plain language in an easy-to-understand manner is important for patients with low functional HL. For example, the term “droplet infection” as a medical term is difficult to understand, however explaining “it is an infection that occurs when the virus is scattered from the mouth when talking” is easier to understand. In addition, providing visual aids to communicate recommended preventive measures, such as wearing masks, will encourage preventive behaviours. Second, for patients with excessively high critical HL, we should consider the positive aspect that caution provides opportunities for appropriate preventive actions. Therefore, dialysis physicians must support patients by identifying misleading information, false claims, and fake news (4). For example, although measures against COVID-19 from reliable information sources (e.g. government or medical institutions) are likely to be beneficial, a claim by a Japanese academic virologist about the risk of miscarriage owing to COVID-19 vaccination via social media may have been difficult to discern as false without comparing different sources or checking for evidence of its origin. Finally, doctors need to be aware of the kinds of patients who are likely to have low functional or high critical HL. In this study, low functional HL was associated with a high number of comorbidities, low educational level, and shorter dialysis duration. In contrast, high critical HL was associated with younger age, low number of comorbidities, and high education level. Thus, addressing individual HL is important because it is affected by several common factors such as education level and comorbidities.

The strengths of this study include the relatively high participation rate and multi- centre design, making the results highly generalisable. In addition, we analysed the association between multidimensional HL and fear of COVID-19 after adjusting for relevant confounding factors. Nonetheless, this study has some limitations. First, although this study measured general HL, it did not directly assess HL related to COVID-19. Second, because this was a cross-sectional study, a possibility of reverse causation exists. For example, it is possible that patients were fearful of COVID-19 and, therefore, were sceptical of health information, including correct information. Third, because most patients had already been vaccinated, we were unable to compare their HL or fear of COVID-19 with those who had not been vaccinated.

In conclusion, fear of COVID-19 among adult Japanese patients undergoing haemodialysis after the widespread COVID-19 vaccine was lower than that in the general population at the beginning of the pandemic. The fear of COVID-19 in patients with low- functional HL can be reduced by providing health information in an easy-to-understand manner. Consequently, honest explanations by healthcare providers may be important for patients with scepticism about correcting information and a strong fear of excessive critical HL. This study is novel in that it investigated the degree of fear of COVID-19 following vaccination and the independent influence of multidimensional HL on fear. An understanding of individual HL may help them to provide appropriate anxiety reduction measures for future pandemics.

## Data availability statement

The datasets generated and/or analysed in the current study are available from the corresponding author upon reasonable request.

## Data Availability

All data produced in the present study are available upon reasonable request to the authors

## Acknowledgements

The authors greatly thank the following researchers, research assistants, and medical staff members for their assistance in collecting the questionnaire-based and clinical information used in this study: Ms. Aki Tairaku (Shin-Yurigaoka General Hospital, Kawasaki-City, Kanagawa); Ms. Takako Saruwatari and Ms. Akiko Kamimura (Kyushu University of Health and Welfare, Nobeoka-City, Miyazaki); Tetuo Ueki, MD, Akio Munakata, MD, Yoshihiko Watanabe, MD (Munakata Clinic, Mobara-City, Chiba); Ms. Yayoi Takanashi, Reiji Masaki, NP, Tomohiko Inoue, MD, Shinnosuke Sugihara, MD, Kanako Nagaoka, MD and Hiroshi Kuji, MD (Kameda Medical Center, Kamogawa-City, Chiba); Kenji Yamaguchi, MD (Awa Regional Medical Center, Tateyama-City, Chiba); Ms. Miyuki Sato (Fukushima Medical University Hospital, Fukushima-City, Fukushima).

## Authors‘ contributions

Research idea and study design: AK, MU, and NK; data acquisition: AK, RI, MU, YK, T. Toishi, MM, TO, YM, and TS; data analysis/interpretation: AK, RI, MU, TS, and NK; statistical analysis: MU, TS, and NK; supervision or mentorship: TO, TS, and NK. Each author contributed important intellectual content during manuscript drafting or revision, agreed to be personally accountable for the individual’s contributions, and ensured that questions on the accuracy or integrity of any portion of the work, even one in which the author was not directly involved, were appropriately investigated and resolved, including documentation in the literature, if appropriate.

